# A DATA-DRIVEN EXAMINATION OF APATHY AND DEPRESSIVE SYMPTOMS IN DEMENTIA WITH INDEPENDENT REPLICATION

**DOI:** 10.1101/2022.09.30.22280551

**Authors:** Miguel Vasconcelos Da Silva, G.J. Melendez-Torres, Zahinoor Ismail, Ingelin Testad, Clive Ballard, Byron Creese, the Alzheimer’s Disease Neuroimaging Initiative

## Abstract

**Introduction:** Apathy is one of the most common neuropsychiatric symptoms (NPS) and is associated with poor clinical outcomes. Research that helps define the apathy phenotype is urgently needed, particularly for clinical and biomarker studies.

**Methods:** We used latent class analysis (LCA) with two independent cohorts to understand how apathy and depression symptoms co-occur statistically. We further explored the relationship between latent class membership, demographics and the presence of other NPS.

**Results:** The LCA identified a 4-class solution (No Symptoms, Apathy, Depression, and Combined Apathy/Depression), reproducible over both cohorts, providing robust support for an apathy syndrome distinct from depression and confirming that an apathy/depression syndrome exists.

**Discussion:** Using a data-driven method, we show distinct and statistically meaningful co-occurrence of apathy and depressive symptoms. There was evidence that these classes have different clinical associations which may help inform diagnostic categories for research studies and clinical practice.

## 1. BACKGROUND

Apathy is one of the most common neuropsychiatric symptoms (NPS) seen in dementia [1], with a reported prevalence of 43% [2] (though estimates vary widely according to dementia stage and study setting). Characterised by a lack of motivation, decreased initiative, akinesia, and emotional indifference [3-7], apathy is a multidimensional syndrome that can lead to functional impairment, poorer treatment response, and greater mortality [8-10]. Understanding the clinical presentation of such a common and clinically important syndrome is of utmost importance both for the effective targeting of new interventions and for the identification of biomarkers or other research aimed at understanding biological correlates.

One of the key diagnostic challenges in apathy is its relationship with depression. Although clinically distinct, symptoms of apathy and of depression are commonly comorbid and have overlapping features [3, 11], thus complicating clinical distinctions between the two [1, 7, 12]. Recognizing this, new diagnostic criteria for apathy in neurocognitive disorders were developed in 2021, representing a major step forward in clinical management of apathy and in associated research by providing a standardised framework for assessment [13]. Central to these criteria is that the symptoms of apathy cannot be explained by another psychiatric condition (e.g., depression), but it remains the case that criteria for apathy can, in principle, be met in the presence of heterogeneous depressive symptomatology. This co-occurrence has the potential to cause a lack of reproducibility, particularly in disease mechanism research where control groups must be precisely defined. It may also be a barrier to targeting the right interventions to the right people. A data-driven approach to distinguish apathy and depression symptomatology can support more precision approaches to treatment and augment the application of the clinically informed diagnostic criteria.

Latent class analysis (LCA) is one example of a data-driven approach. A form of latent variable modelling, LCA describes combinations of generally binary variables in terms of statistically likely patterns that are not directly observed (i.e., latent classes), where these patterns are characterised on the basis of the conditional probability of each binary variable within a class.

Thus, using LCA on questionnaire-based measures of apathy and depression, we aimed to 1) examine how symptoms co-occur in two independent observational cohort studies; and 2) establish whether the latent classes identified were associated with different NPS profiles.

## 2. METHODS

### 2.1 Sample descriptions

#### 2.1.1 L-study

Participants were from the Measuring Long Term Outcomes in People with Dementia in Care Homes (L-study). This study has been running since 2015 through the King’s College London and Maudsley Care Home Research Network (CHRN). Participants with a diagnosis of dementia living in care homes across the southeast of England were recruited. Consent was obtained from participants or next of kin if participants were unable to consent for themselves. The recruitment and assessments were completed by a trained researcher. The Southampton and South West Hampshire Research Ethics Committee A (formally South Central – Southampton A) approved the study (REC number 13/SC/0265).

#### 2.1.2 ADNI

Additional data used in the preparation of this article were obtained from the Alzheimer’s Disease Neuroimaging Initiative (ADNI) database (adni.loni.usc.edu). The ADNI was launched in 2003 as a public-private partnership, led by Principal Investigator Michael W. Weiner, MD. The primary goal of ADNI has been to test whether serial magnetic resonance imaging (MRI), positron emission tomography (PET), other biological markers, and clinical and neuropsychological assessment can be combined to measure the progression of mild cognitive impairment (MCI) and early Alzheimer’s disease (AD). For up-to-date information, see www.adni-info.org.

### 2.2 Measures

#### 2.2.1 Neuropsychiatric symptoms

NPS were assessed with the Neuropsychiatric Inventory (NPI), completed by a researcher-led interview with an informed caregiver [14]. The NPI assesses the following 12 NPS, within a reference period of the past 4 weeks: delusions; hallucinations; agitation/aggression; depression/dysphoria; anxiety; elation/euphoria; apathy/indifference; disinhibition; irritability/lability; aberrant motor behaviour; sleep and night-time behaviour disorders, and appetite and eating disorders [15]. Each item is rated as present or absent. If a screening question is rated present, a number of sub-questions are then completed which cover detailed NPS relating to each domain; each is coded with a yes or no response. If any of these symptoms are rated present, frequency (1-4) and severity (1-3) are rated, with the product of these representing the overall symptom score.

For the L-study, the NPI Nursing Home (NPI-NH) version was used as this is the adapted version for institutional settings whilst the ADNI cohort used the standard NPI [16].

#### 2.2.2 Apathy and Depression

Apathy and depression were measured using the sub-questions on item G (Apathy) and item D (Depression) of the NPI (see Appendix A for list of questions). For the NPI-NH there are 15 sub-questions while for the standard NPI there are 16, with the standard NPI scale used in ADNI having one fewer sub-question on the Apathy section.

#### 2.2.3 Other neuropsychiatric symptoms

Our second aim was to evaluate the relationship between latent classes of apathy and depression and other NPS. To do this, NPS status (defined as present or absent) was determined for the remaining 10 items on the NPI using binary coding. Participants with a frequency x severity score of ≥1 were considered as ‘symptom present’. The symptom was considered absent if the score was 0.

#### 2.2.4 Dementia severity

The Clinical Dementia Rating (CDR) is a scale for assessing, diagnosing and staging cognitive impairment [17]. A researcher/clinician conducts a semi-structured interview with the patient and a reliable informant to rate performance in six domains: memory; orientation; judgement and problem solving; community affairs, homes and hobbies; and personal care. The domains are rated in terms of the impairment level (0 = none, 0.5 = questionable, 1 = mild, 2 = moderate, 3 = severe). A final global rating is then calculated to indicate the level of impairment (0 = normal cognition, 0.5 = questionable or very mild dementia, 1 = mild dementia, 2 = moderate dementia, and 3 = severe dementia).

### 2.3 Data Cleaning

Participants with a CDR of 0 or 0.5 were excluded from both cohorts given this would indicate normal cognition or questionable dementia. Data were checked for completeness, participants with missing age, sex, or CDR were removed from the dataset. Participants with missing NPI/NPI-NH items D and G were also excluded. A total of 22 participants were removed from L-study cohort and a total of 135 participants were removed from ADNI. Furthermore, within the ADNI cohort only 4 participants had a CDR of 3 so these were merged with the group scoring 2. See Appendix B for study flow diagram.

### 2.4 Statistical Analysis

To summarise, first we estimated latent classes of the NPI apathy and depression subscale questions with two to six class solutions. This was followed by the bootstrap test to analyse the best fitting solution. Then we explored the relationship between latent classes from the best fitting LCA model and the presence of other NPI domains. The same steps were performed for the L-study and ADNI datasets. Further detail is provided below.

We used the Stata LCA plugin version 1.2.1 [18] to estimate the latent classes, measured by categorical indicators, within the NPI category D (depression) and G (apathy) sub-questions. The LCA Bootstrap version 1.0 plugin [19] was then used to evaluate models with two to six latent classes and determine the optimal number of classes using the model-fit criteria of bootstrapped likelihood ratio test (BLRT), adjusted Bayesian Information Criterion (aBIC), scaled relative entropy, and log likelihood. A simulation study has shown that BLRT was the best test to identify the correct number of classes, followed by the Bayesian Information Criterion and the aBIC [20]. Thus, a model with k classes would be considered a better fit than a model with k-1 classes if it had a lower BLRT, p-value and a lower aBIC value. In addition, we considered the clinical interpretability of the final model.

Following this we used the plugin LCA_Distal_BCH version 1.1 [21] to analyse the relationship between the latent classes with other NPI items. This test estimates the association between a latent class variable and an observed distal outcome, i.e., other NPI items, using the approach of Bolck, Croon, and Hagenaars [22], as adapted by Vermunt [23], allowing for the probability of misclassification of classes. Distal probabilities for each NPI item for each latent class are reported along with the Wald test for significance. The Wald test tests the null hypothesis that the probabilities are the same across all latent classes. Pairwise comparisons between latent classes were then performed to determine which latent classes differed with respect to the probabilities of other NPI items.

## 3. RESULTS

### 3.1 Participant characteristics

After data cleaning, the L-study comprised of 326 people. Mean age was 87 (SD 6.99), 73% were female and median CDR was 2 (see Table 1). The ADNI data comprised of 271 people. Mean age was 74 (SD 7.4), 42% were female with a median CDR of 1 (see Table 1).

**Table 1.**
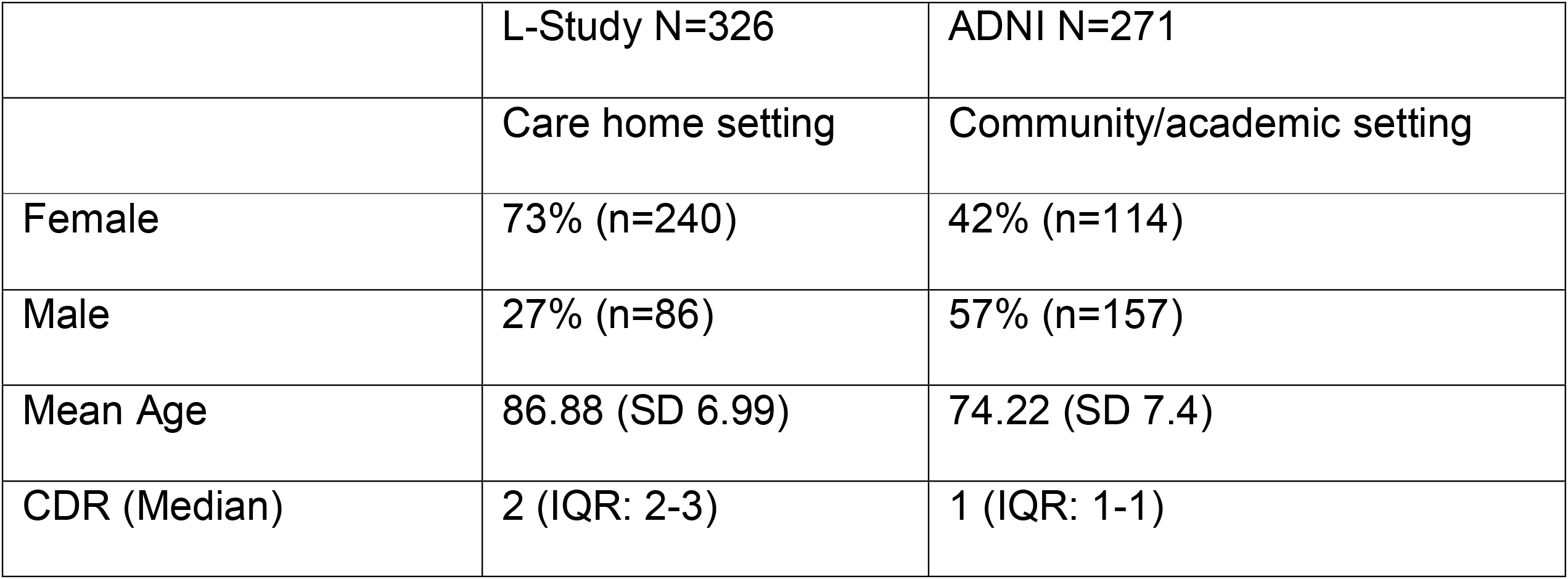
Cohorts Demographics.

### 3.2 Latent Class analysis identifies a reproducible 4-class solution

#### 3.2.1 L-study

A 4-class model was considered optimal based on the higher aBIC score and higher log likelihood score, p-value and clinical interpretability. Although the 5-class model had same p-value, on balance comparing the other metrics and clinical interpretability the 4-class model was chosen (see Table 2).

**Table 2.**
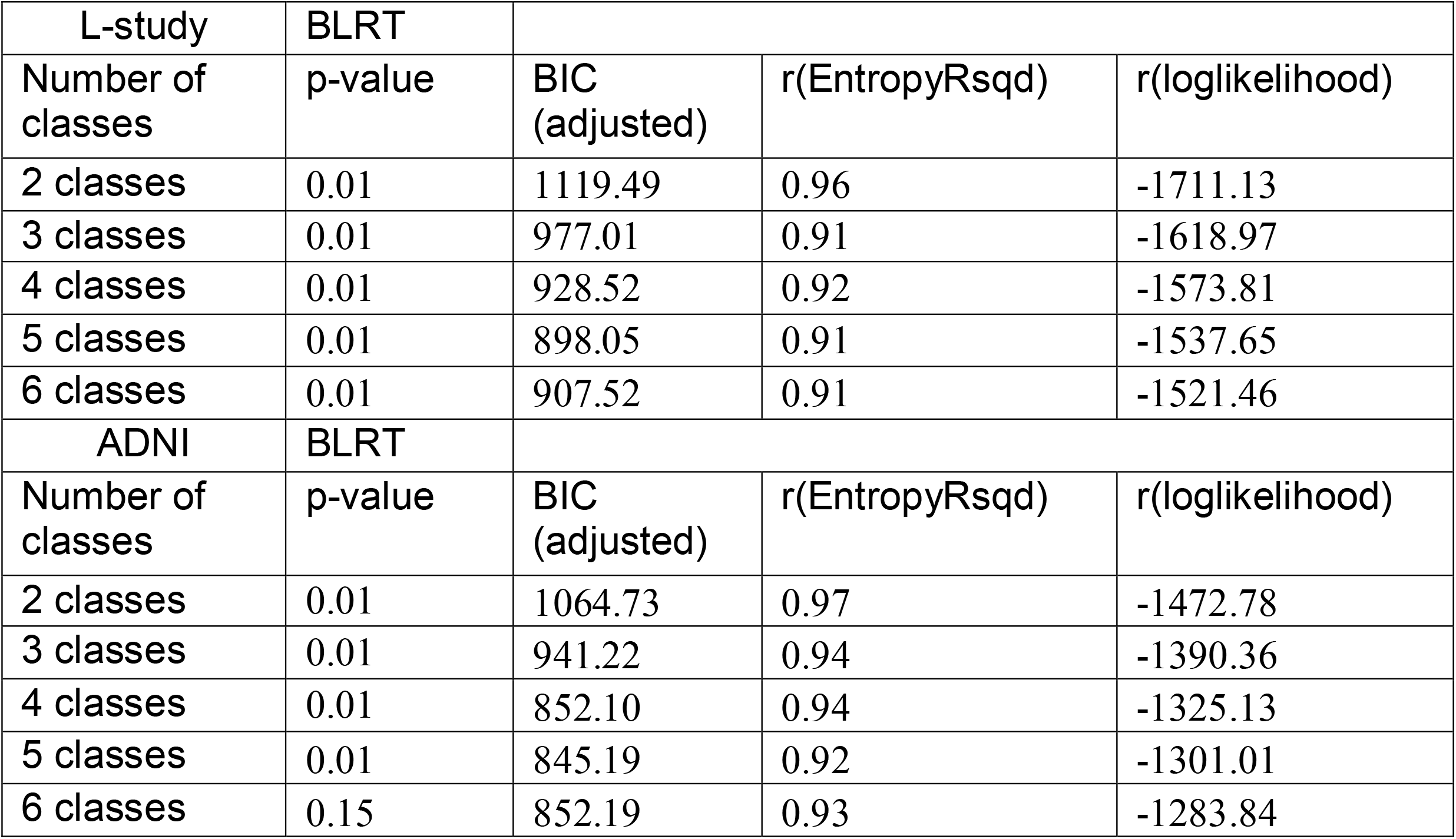
Bootstrap Likelihood Ratio test and Scalars for best model fit.

The four classes were labelled No Symptoms, Combined Apathy/Depression, Depression, and Apathy. Looking at the symptom probability across the different classes (Figure 1), the No Symptoms class showed very low or zero probability of apathy and depressive symptoms; the Combined Apathy/Depression class exhibited high probabilities across a range of depressive and apathy symptoms; the Depression class comprised mostly of depressive symptoms with low probability of some apathy symptoms; the Apathy class had high probabilities across apathy symptoms and very low or no probability of depressive symptoms. The probabilities of participants falling into each class was 42.13%, 18.37%, 9.06% and 30.42% for the No Symptom, Combined Apathy/Depression, Depression, and Apathy classes respectively (see Appendix C).

**Figure 1.**
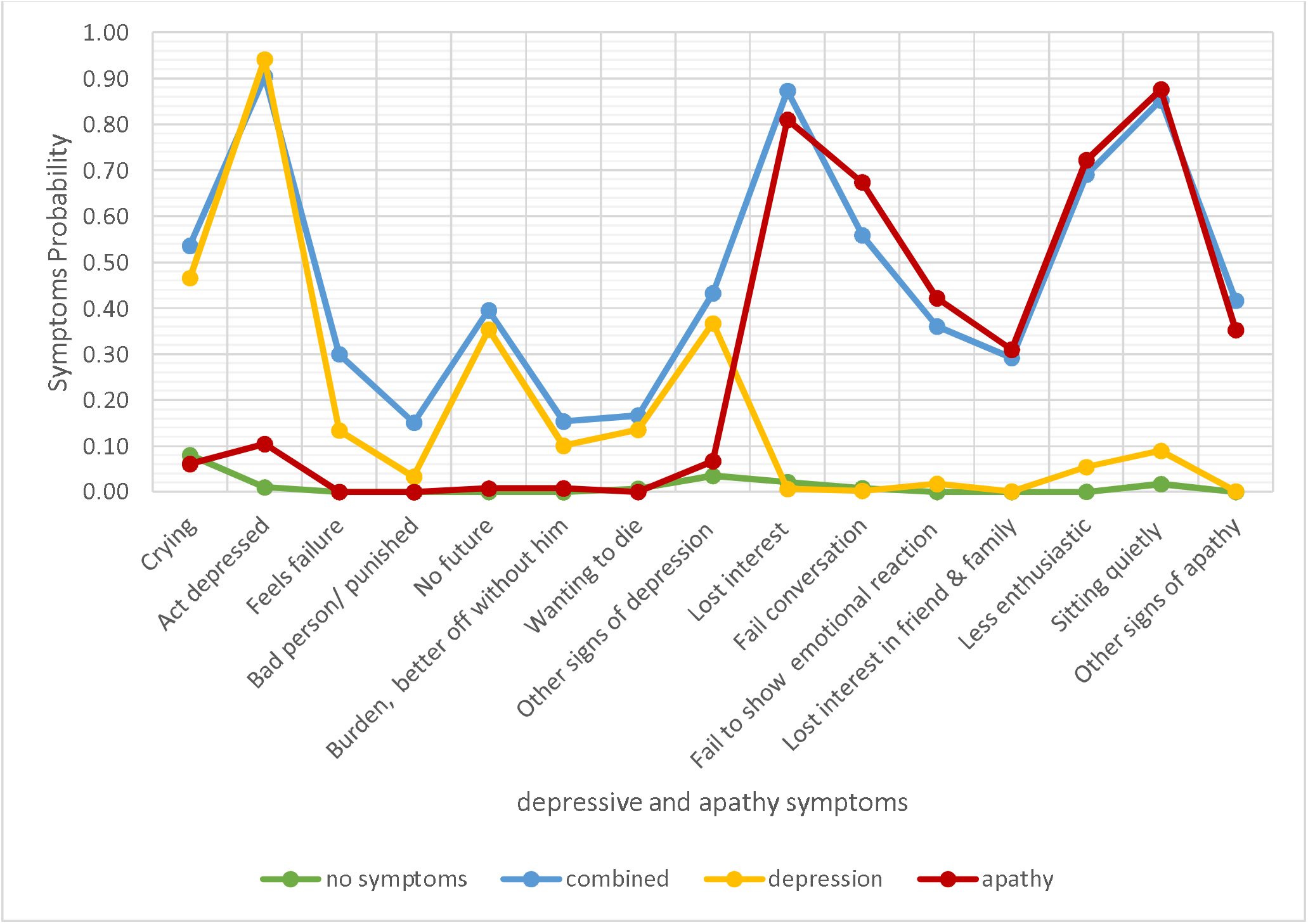
Apathy and depression symptoms probability across the 4 classes in L-study.

#### 3.2.2 ADNI

Considering aBIC and p-value and clinical interpretability, a 4-class model was again considered optimal, with a notable similarity to the L-study across the symptom probabilities (Figure 2). We note that a case could be made for selecting a 5-class model, however the additional class in the 5-class model was very similar to class 2 in the 4-class model (see Appendix E for more detailed discussion of model selection).

**Figure 2.**
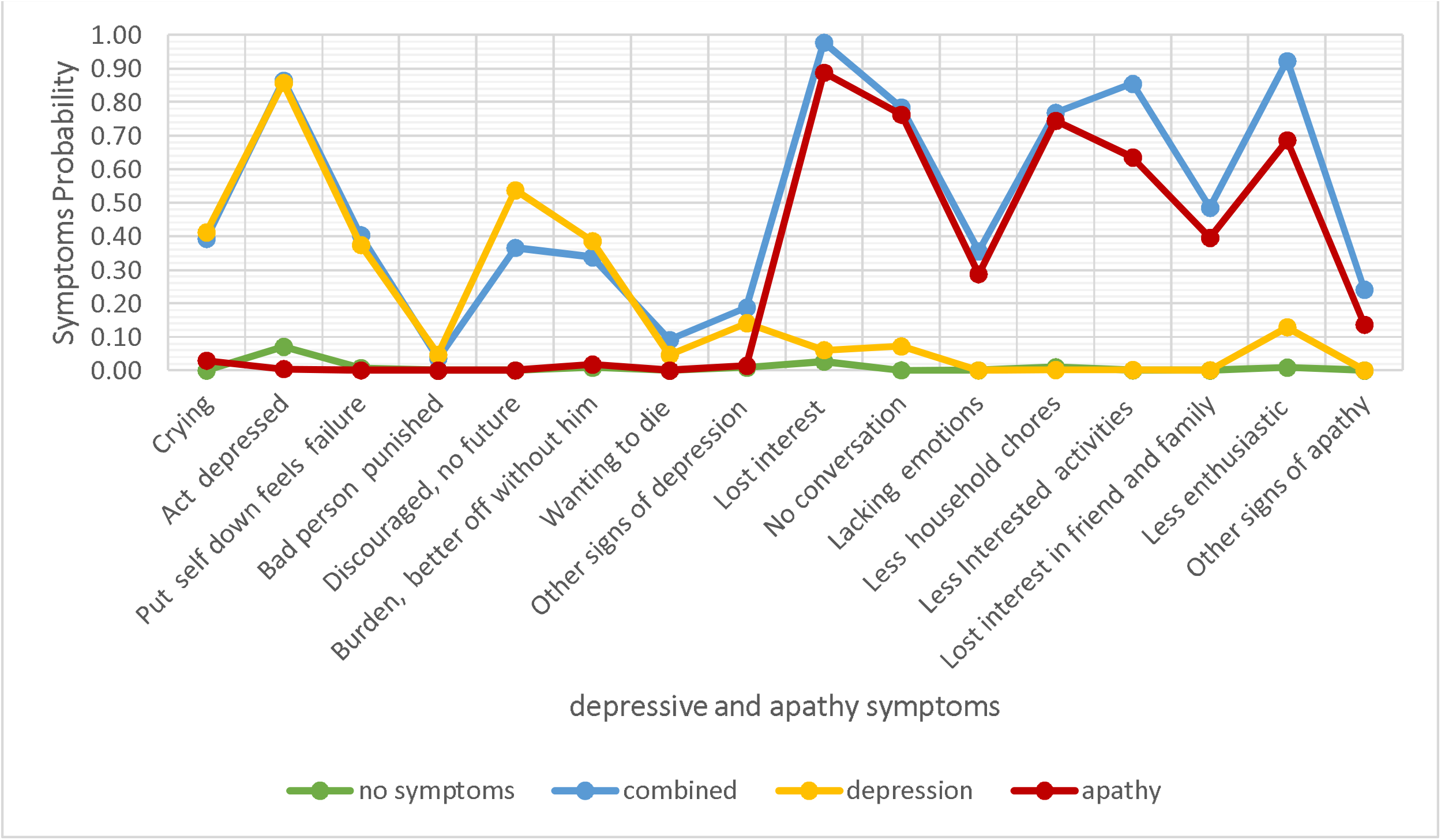
Apathy and depression symptoms probability across the 4 classes in ADNI.

The four classes were given the same labels as the L-Study, with the same descriptions. The probabilities of participants falling into each class in the 4-class model were 40.7%, 20.1%, 15.6% and 23.6% for the No Symptoms, Combined Apathy/Depression, Depression, and Apathy classes respectively (see Appendix C).

### 3.3 In individuals with apathy, co-morbid depressive symptoms have higher probability of additional NPS burden

Broadly, the latent classes that were characterised by the presence of depressive symptoms (i.e., Combined Apathy/Depression and Depression) were associated with a higher burden of comorbid NPS, particularly delusions, anxiety and irritability, which replicated across the two cohorts. The Apathy class (i.e., no depression symptoms) relative to the No Symptoms class was associated with a higher probability for agitation in the L-study, but not in ADNI. This was the only comparison where Apathy conferred a higher probability for any comorbid NPS.

Specific pairwise comparisons between latent classes are described in more detail below. Only NPS domains where there was evidence of replication across the two cohorts are considered in detail and shown in Table 3. All other results are in Table F.1. in Appendix F.

**Table 3.**
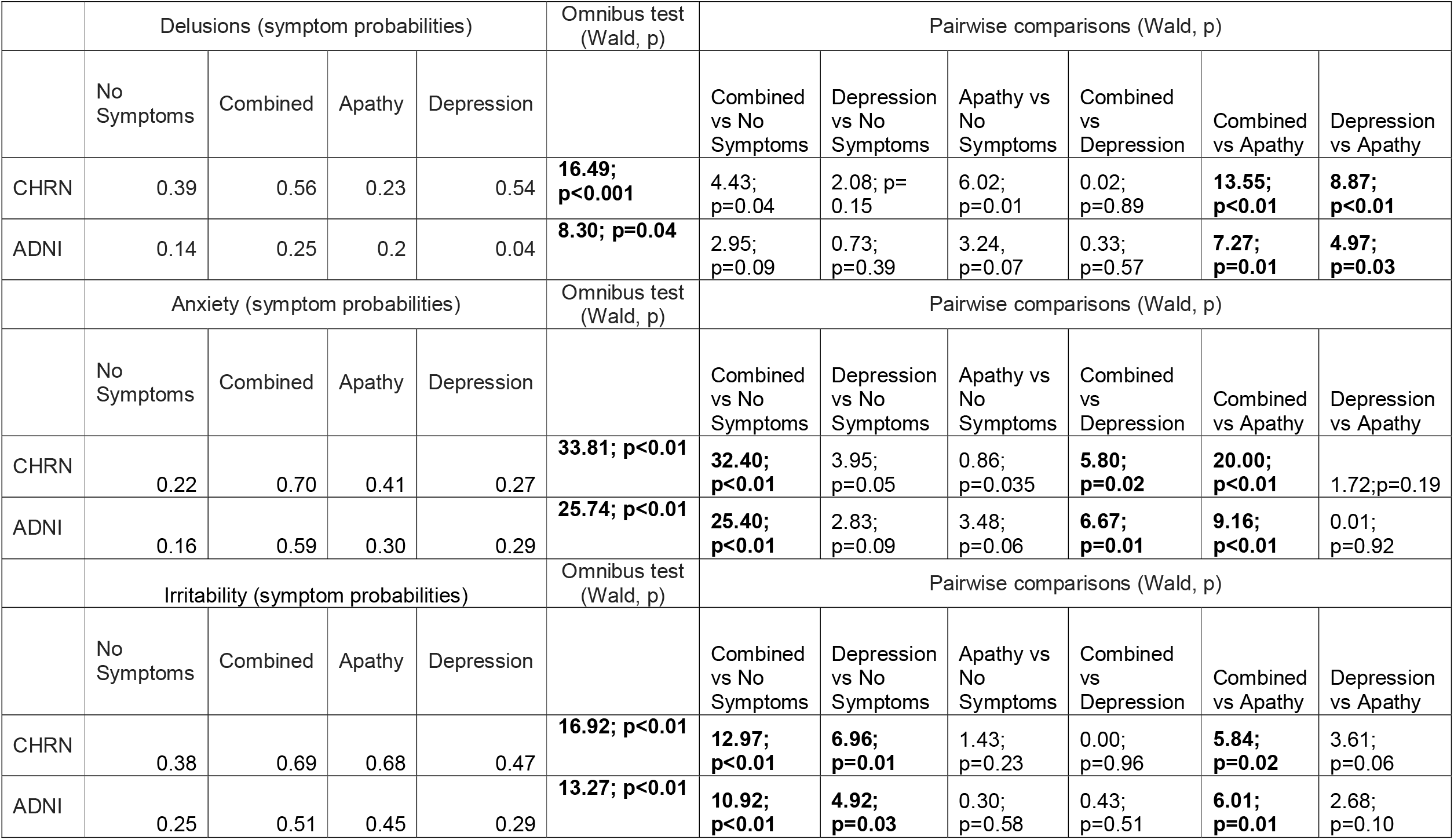
Delusions, Anxiety and irritability probabilities across the 4 classes, with pairwise comparisons.

#### 3.3.1 Delusions

The probability for delusions was highest in the classes characterised by depressive symptoms (Combined Apathy/Depression class in L-study = 0.56 and in ADNI =0.25; Depression class in L-study = 0.54 and in ADNI = 0.20) and lowest in the No Symptoms and Apathy class (No symptoms L-study= 0.39 ADNI = 0.14; Apathy class L-study=0.23; ADNI=0.04), with L-study Wald test= 16.49, p<0.001 and ADNI Wald test=8.3, p=0.04 see Table 3.

#### 3.3.2 Anxiety

The probability for Anxiety was highest in the classes characterised by depressive symptoms (Combined Apathy/Depression class L-study = 0.70 and in ADNI =0.59; Depression class in L-study = 0.41 and in ADNI = 0.30) and lowest in the No Symptoms and Apathy class (No symptoms in L-study= 0.22 ADNI = 0.16; Apathy class in L-study=0.27 ADNI=0.29], with L-study Wald test= 33.81, p<0.001 and ADNI Wald test=25.74, p<0.001 see Table 3.

#### 3.3.3 Irritability

The probability for irritability was highest in the classes characterised by depressive symptoms (Combined Apathy/Depression class L-study = 0.69 and in ADNI =0.51; Depression class in L-study = 0.68 and in ADNI = 0.45) and lowest in the No Symptoms and Apathy class (No symptoms in L-study= 0.38 ADNI = 0.25; Apathy class in L-study=0.47 ADNI=0.29], with L-study Wald test= 16.92, p<0.001 and ADNI Wald test=13.27, p=0.004 see Table 3.

#### 3.3.4 Other associations

Other associations that did not replicate across the two datasets are shown in the supplement, but we highlight here the findings relating to agitation as one of the most clinically interesting of the remaining findings. The Combined Apathy/Depression and Apathy classes had a higher probability of agitation relative to the No Symptoms class in the L-study only (probability of agitation for No Symptoms, Combined Apathy/Depression, Depression, and Apathy Class was 0.52, 0.80, 0.64, 0.72 respectively with overall Wald Test L-study= 17.04, p<0.01). This did not replicate in ADNI (probability of agitation for No Symptoms, Combined Apathy/Depression, Depression, and Apathy Class was 0.25, 0.4, 0.4 and 0.39 respectively with overall Wald Test 5.53, p=0.13) however the direction of effect was similar.

## 4. DISCUSSION

Here, we used LCA to delineate symptoms of apathy and depression in dementia. The reproducible 4-class solution provides robust data-driven validation of an apathy syndrome that is distinct from depression and a combined apathy/depression syndrome. This conclusion is supported by our analysis of neuropsychiatric symptom associations with each of the four classes where we show that apathy is generally only associated with a more severe NPS profile when co-morbid depressive symptoms are present.

The broad implication of these findings is that in apathy research, depression should also be routinely measured and individuals with comorbid symptoms may have to be considered a separate group to those with a distinct apathy syndrome. This could explain some of the heterogeneity observed in NPS associations in other studies of apathy. For example, one study observed that apathy was associated with higher NPI scores across a range of NPS, including delusions, irritability and anxiety [24]. In light of the findings from the present study, it is possible that the reported associations in this previous study were being driven by the presence of comorbid depressive symptoms in the apathy group. Future studies measuring clinical and biological correlates should consider the possibility of comorbid depressive symptoms as a confounder in order to produce reproducible research.

The latest diagnostic criteria for apathy stipulate that apathy cannot be due to another illness, disability, or substance abuse but are silent on accompanying neuropsychiatric features. The existence of the Combined Apathy/Depression class suggests further consideration of depressive symptoms (which may not meet criteria for clinical depression) as an accompanying feature of apathy.

In terms of other NPS burden, there was a striking consistency for a higher probability of delusions, anxiety, and irritability in the two classes that captured depressive symptoms (i.e., the Depression and Combined Apathy/Depression classes) relative to the No Symptoms and Apathy classes. This pattern of association points to an affective syndrome with psychotic features, validating both the focus on the relationship between these syndromes in the new IPA and ISTAART criteria for psychosis in neurocognitive disorders [25, 26], and results from previous studies reporting the co-occurrence of both syndromes [27-30]. Mechanistically, a recent genome wide association study observed a positive association between genetic risk for depressive symptoms and Alzheimer’s disease with psychosis, while in younger-aged samples genetic correlations between irritability and depressive symptoms have been observed [31, 32]. These observations, alongside the current findings, reflect current opinion that comorbid behavioural symptoms may impact response to treatments [33].

Another interesting observation is that there was a higher probability for agitation in the Apathy class relative to the No Symptoms class in the L-study. Although only present in one cohort, in which mean age was higher and median dementia severity was slightly greater, this was the only instance where the Apathy class was associated with a higher probability of another NPS. The lack of replication may be due to the more advanced dementia seen in the L-study, which is a care home study, where we would expect agitation to be more common [34]. Nonetheless, this may be a clinically relevant finding. The association of apathy with comorbid agitation is an important relationship to explore, as these individuals may require additional support and are often referred for specialist consultation. We would argue this is a relationship worth examining more closely in future research to determine whether agitation should be considered an accompanying feature of apathy and incorporated into later revision of the diagnostic criteria (which is currently silent on comorbid NPS). A clear path to take these findings forward is to perform LCA on apathy and depressive questionnaire responses and map the classes identified onto independently rated apathy diagnostic criteria in the same individuals. This analysis would provide the necessary data to evaluate the extent to which application of the criteria captures individuals with depressive symptoms.

A key strength of this study is the replication in two independent cohorts which confers a high degree of confidence in our findings. This is important as caregiver reports may be influenced by caregiver mood, cultural beliefs, and the educational level of the caregiver [35], although also the carer information has the advantage of not being influenced by anosognosia. However, differences in the dementia severity and demographics between the ADNI and the L-study mean we must acknowledge the possibility that non-replication of neuropsychiatric symptom associations is due to these differences (as discussed above with respect to agitation). We also acknowledge the inherent limitation of the NPI in measuring apathy, particularly the relative weighting given to items capturing diminished interest (with a lesser focus on the emotional and initiative dimensions). However, if the latent classes captured in this study reflect true unobserved and mutually exclusive groupings, then these should be stable across any measures of depression and apathy, provided appropriate questionnaire items are measured. This is a hypothesis that can readily be tested by future research in other samples with different NPS measurements.

In summary, this study supports the existence four latent classes (No Symptoms, Combined Apathy/Depression, Depression and Apathy) replicated in two independent cohorts. In analysis of comorbid NPS burden associated with these classes, we show that the presence of depressive symptoms is the major driver. These findings do not compete with the diagnostic criteria for apathy; rather they should be considered supporting evidence for future iterations, potentially qualifying comorbid NPS symptoms or syndromes. Specifically, we propose that future consideration be given to depressive and agitation symptoms as accompanying neuropsychiatric features of apathy.

## Data Availability

Data is available from ADNI. L-study data is not yet publically available.

## APPENDIX

**The following are the published list of collaborators associated with ADNI**

Michael Weiner, MD (UC San Francisco, Principal Investigator, Executive Committee); Paul Aisen, MD (UC San Diego, ADCS PI and Director of Coordinating Center Clinical Core, Executive Committee, Clinical Core Leaders); Ronald Petersen, MD, PhD (Mayo Clinic,Rochester, Executive Committee, Clinical Core Leader); Clifford R. Jack, Jr., MD (Mayo Clinic, Rochester, Executive Committee, MRI Core Leader); William Jagust, MD (UC Berkeley, Executive Committee; PET Core Leader); John Q. Trojanowki, MD, PhD (U Pennsylvania, Executive Committee, Biomarkers Core Leader); Arthur W. Toga, PhD (USC, Executive Committee, Informatics Core Leader); Laurel Beckett, PhD (UC Davis, Executive Committee, Biostatistics Core Leader); Robert C. Green, MD, MPH (Brigham and Women’s Hospital, Harvard Medical School, Executive Committee and Chair of Data and Publication Committee); Andrew J. Saykin, PsyD (Indiana University, Executive Committee, Genetics Core Leader); John Morris, MD (Washington University St. Louis, Executive Committee, Neuropathology Core Leader); Leslie M. Shaw (University of Pennsylvania, Executive Committee, Biomarkers Core Leader); Enchi Liu, PhD (Janssen Alzheimer Immunotherapy, ADNI 2 Private Partner Scientific Board Chair); Tom Montine, MD, PhD (University of Washington); Ronald G. Thomas, PhD (UC San Diego); Michael Donohue, PhD (UC San Diego); Sarah Walter, MSc (UC San Diego); Devon Gessert (UC San Diego); Tamie Sather, MS (UC San Diego,); Gus Jiminez, MBS (UC San Diego); Danielle Harvey, PhD (UC Davis;); Michael Donohue, PhD (UC San Diego); Matthew Bernstein, PhD (Mayo Clinic, Rochester); Nick Fox, MD (University of London); Paul Thompson, PhD (USC School of Medicine); Norbert Schuff, PhD (UCSF MRI); Charles DeCArli, MD (UC Davis); Bret Borowski, RT (Mayo Clinic); Jeff Gunter, PhD (Mayo Clinic); Matt Senjem, MS (Mayo Clinic); Prashanthi Vemuri, PhD (Mayo Clinic); David Jones, MD (Mayo Clinic); Kejal Kantarci (Mayo Clinic); Chad Ward (Mayo Clinic); Robert A. Koeppe, PhD (University of Michigan, PET Core Leader); Norm Foster, MD (University of Utah); Eric M. Reiman, MD (Banner Alzheimer’s Institute); Kewei Chen, PhD (Banner Alzheimer’s Institute); Chet Mathis, MD (University of Pittsburgh); Susan Landau, PhD (UC Berkeley); Nigel J. Cairns, PhD, MRCPath (Washington University St. Louis); Erin Householder (Washington University St. Louis); Lisa Taylor Reinwald, BA, HTL (Washington University St. Louis); Virginia Lee, PhD, MBA (UPenn School of Medicine); Magdalena Korecka, PhD (UPenn School of Medicine); Michal Figurski, PhD (UPenn School of Medicine); Karen Crawford (USC); Scott Neu, PhD (USC); Tatiana M. Foroud, PhD (Indiana University); Steven Potkin, MD UC (UC Irvine); Li Shen, PhD (Indiana University); Faber Kelley, MS, CCRC (Indiana University); Sungeun Kim, PhD (Indiana University); Kwangsik Nho, PhD (Indiana University); Zaven Kachaturian, PhD (Khachaturian, Radebaugh & Associates, Inc and Alzheimer’s Association’s Ronald and Nancy Reagan’s Research Institute); Richard Frank, MD, PhD (General Electric); Peter J. Snyder, PhD (Brown University); Susan Molchan, PhD (National Institute on Aging/ National Institutes of Health); Jeffrey Kaye, MD (Oregon Health and Science University); Joseph Quinn, MD (Oregon Health and Science University); Betty Lind, BS (Oregon Health and Science University); Raina Carter, BA (Oregon Health and Science University); Sara Dolen, BS (Oregon Health and Science University); Lon S. Schneider, MD (University of Southern CaliforGroups Acknowledgements Journal Format nia); Sonia Pawluczyk, MD (University of Southern California); Mauricio Beccera, BS University of Southern California); Liberty Teodoro, RN (University of Southern California); Bryan M. Spann, DO, PhD (University of Southern California); James Brewer, MD, PhD (University of California San Diego); Helen Vanderswag, RN (University of California San Diego); Adam Fleisher, MD (University of California San Diego); Judith L. Heidebrink, MD, MS (University of Michigan); Joanne L. Lord, LPN, BA, CCRC (University of Michigan); Ronald Petersen, MD, PhD (Mayo Clinic, Rochester); Sara S. Mason, RN (Mayo Clinic, Rochester); Colleen S. Albers, RN (Mayo Clinic, Rochester); David Knopman, MD (Mayo Clinic, Rochester); Kris Johnson, RN (Mayo Clinic, Rochester); Rachelle S. Doody, MD, PhD (Baylor College of Medicine); Javier Villanueva Meyer, MD (Baylor College of Medicine); Munir Chowdhury, MBBS, MS (Baylor College of Medicine); Susan Rountree, MD (Baylor College of Medicine); Mimi Dang, MD (Baylor College of Medicine); Yaakov Stern, PhD (Columbia University Medical Center); Lawrence S. Honig, MD, PhD (Columbia University Medical Center); Karen L. Bell, MD (Columbia University Medical Center); Beau Ances, MD (Washington University, St. Louis); John C. Morris, MD (Washington University, St. Louis); Maria Carroll, RN, MSN (Washington University, St. Louis); Sue Leon, RN, MSN (Washington University, St. Louis); Erin Householder, MS, CCRP (Washington University, St. Louis); Mark A. Mintun, MD (Washington University, St. Louis); Stacy Schneider, APRN, BC, GNP (Washington University, St. Louis); Angela Oliver, RN, BSN, MSG; Daniel Marson, JD, PhD (University of Alabama Birmingham); Randall Griffith, PhD, ABPP (University of Alabama Birmingham); David Clark, MD (University of Alabama Birmingham); David Geldmacher, MD (University of Alabama Birmingham); John Brockington, MD (University of Alabama Birmingham); Erik Roberson, MD (University of Alabama Birmingham); Hillel Grossman, MD (Mount Sinai School of Medicine); Effie Mitsis, PhD (Mount Sinai School of Medicine); Leyla deToledoMorrell, PhD (Rush University Medical Center); Raj C. Shah, MD (Rush University Medical Center); Ranjan Duara, MD (Wien Center); Daniel Varon, MD (Wien Center); Maria T. Greig, HP (Wien Center); Peggy Roberts, CNA (Wien Center); Marilyn Albert, PhD (Johns Hopkins University); Chiadi Onyike, MD (Johns Hopkins University); Daniel D’Agostino II, BS (Johns Hopkins University); Stephanie Kielb, BS (Johns Hopkins University); James E. Galvin, MD, MPH (New York University); Dana M. Pogorelec (New York University); Brittany Cerbone (New York University); Christina A. Michel (New York University); Henry Rusinek, PhD (New York University); Mony J de Leon, EdD (New York University); Lidia Glodzik, MD, PhD (New York University); Susan De Santi, PhD (New York University); P. Murali Doraiswamy, MD (Duke University Medical Center); Jeffrey R. Petrella, MD (Duke University Medical Center); Terence Z. Wong, MD (Duke University Medical Center); Steven E. Arnold, MD (University of Pennsylvania); Jason H. Karlawish, MD (University of Pennsylvania); David Wolk, MD (University of Pennsylvania); Charles D. Smith, MD (University of Kentucky); Greg Jicha, MD (University of Kentucky); Peter Hardy, PhD (University of Kentucky); Partha Sinha, PhD (University of Kentucky); Elizabeth Oates, MD (University of Kentucky); Gary Conrad, MD (University of Kentucky); Oscar L. Lopez, MD (University of Pittsburgh); MaryAnn Oakley, MA (University of Pittsburgh); Donna M. Simpson, CRNP, MPH (University of Pittsburgh); Anton P. Porsteinsson, MD (University of Rochester Medical Center); Bonnie S. Goldstein, MS, NP (University of Rochester Medical Center); Kim Martin, RN (University of Rochester Medical Center); Kelly M. Makino, BS (University of Rochester Medical Center); M. Saleem Ismail, MD (University of Rochester Medical Center); Connie Brand, RN (University of Rochester Medical Center); Ruth A. Mulnard, DNSc, RN, FAAN (University of California, Irvine); Gaby Thai, MD (University of California, Irvine); Catherine Mc Adams Ortiz, MSN, RN, A/GNP (University of California, Irvine); Kyle Womack, MD (University of Texas Southwestern Medical School); Dana Mathews, MD, PhD (University of Texas Southwestern Medical School); Mary Quiceno, MD (University of Texas Southwestern Medical School); Ramon Diaz Arrastia, MD, PhD (University of Texas Southwestern Medical School); Richard King, MD (University of Texas Southwestern Medical School); Myron Weiner, MD (University of Texas Southwestern Medical School); Kristen Martin Cook, MA (University of Texas Southwestern Medical School); Michael DeVous, PhD (University of Texas Southwestern Medical School); Allan I. Levey, MD, PhD (Emory University); James J. Lah, MD, PhD (Emory University); Janet S. Cellar, DNP, PMHCNS BC (Emory University); Jeffrey M. Burns, MD (University of Kansas, Medical Center); Heather S. Anderson, MD (University of Kansas, Medical Center); Russell H. Swerdlow, MD (University of Kansas, Medical Center); Liana Apostolova, MD (University of California, Los Angeles); Kathleen Tingus, PhD (University of California, Los Angeles); Ellen Woo, PhD (University of California, Los Angeles); Daniel H.S. Silverman, MD, PhD (University of California, Los Angeles); Po H. Lu, PsyD (University of California, Los Angeles); George Bartzokis, MD (University of California, Los Angeles); Neill R Graff Radford, MBBCH, FRCP (London) (Mayo Clinic, JacksonGroups Acknowledgements Journal Format 2 ville); Francine Parfitt, MSH, CCRC (Mayo Clinic, Jacksonville); Tracy Kendall, BA, CCRP (Mayo Clinic, Jacksonville); Heather Johnson, MLS, CCRP (Mayo Clinic, Jacksonville); Martin R. Farlow, MD (Indiana University); Ann Marie Hake, MD (Indiana University); Brandy R. Matthews, MD (Indiana University); Scott Herring, RN, CCRC (Indiana University); Cynthia Hunt, BS, CCRP (Indiana University); Christopher H. van Dyck, MD (Yale University School of Medicine); Richard E. Carson, PhD (Yale University School of Medicine); Martha G. MacAvoy, PhD (Yale University School of Medicine); Howard Chertkow, MD (McGill Univ., Montreal Jewish General Hospital); Howard Bergman, MD (McGill Univ., Montreal Jewish General Hospital); Chris Hosein, Med (McGill Univ., Montreal Jewish General Hospital); Sandra Black, MD, FRCPC (Sunnybrook Health Sciences, Ontario); Dr Bojana Stefanovic (Sunnybrook Health Sciences, Ontario); Curtis Caldwell, PhD (Sunnybrook Health Sciences, Ontario); Ging Yuek Robin Hsiung, MD, MHSc, FRCPC (U.B.C. Clinic for AD & Related Disorders); Howard Feldman, MD, FRCPC (U.B.C. Clinic for AD & Related Disorders); Benita Mudge, BS (U.B.C. Clinic for AD & Related Disorders); Michele Assaly, MA Past (U.B.C. Clinic for AD & Related Disorders); Andrew Kertesz, MD (Cognitive Neurology St. Joseph’s, Ontario); John Rogers, MD (Cognitive Neurology St. Joseph’s, Ontario); Dick Trost, PhD (Cognitive Neurology St. Joseph’s, Ontario); Charles Bernick, MD (Cleveland Clinic Lou Ruvo Center for Brain Health); Donna Munic, PhD (Cleveland Clinic Lou Ruvo Center for Brain Health); Diana Kerwin, MD (Northwestern University); Marek Marsel Mesulam, MD (Northwestern University); Kristine Lipowski, BA (Northwestern University); Chuang Kuo Wu, MD, PhD (Northwestern University); Nancy Johnson, PhD (Northwestern University); Carl Sadowsky, MD (Premiere Research Inst (Palm Beach Neurology)); Walter Martinez, MD (Premiere Research Inst (Palm Beach Neurology)); Teresa Villena, MD (Premiere Research Inst (Palm Beach Neurology)); Raymond Scott Turner, MD, PhD (Georgetown University Medical Center); Kathleen Johnson, NP (Georgetown University Medical Center); Brigid Reynolds, NP (Georgetown University Medical Center); Reisa A. Sperling, MD (Brigham and Women’s Hospital); Keith A. Johnson, MD (Brigham and Women’s Hospital); Gad Marshall, MD (Brigham and Women’s Hospital); Meghan Frey (Brigham and Women’s Hospital); Jerome Yesavage, MD (Stanford University); Joy L. Taylor, PhD (Stanford University); Barton Lane, MD (Stanford University); Allyson Rosen, PhD (Stanford University); Jared Tinklenberg, MD (Stanford University); Marwan N. Sabbagh, MD (Banner Sun Health Research Institute); Christine M. Belden, PsyD (Banner Sun Health Research Institute); Sandra A. Jacobson, MD (Banner Sun Health Research Institute); Sherye A. Sirrel, MS (Banner Sun Health Research Institute); Neil Kowall, MD (Boston University); Ronald Killiany, PhD (Boston University); Andrew E. Budson, MD (Boston University); Alexander Norbash, MD (Boston University); Patricia Lynn Johnson, BA (Boston University); Thomas O. Obisesan, MD, MPH (Howard University); Saba Wolday, MSc (Howard University); Joanne Allard, PhD (Howard University); Alan Lerner, MD (Case Western Reserve University); Paula Ogrocki, PhD (Case Western Reserve University); Leon Hudson, MPH (Case Western Reserve University); Evan Fletcher, PhD (University of California, Davis Sacramento); Owen Carmichael, PhD (University of California, Davis Sacramento); John Olichney, MD (University of California, Davis Sacramento); Charles DeCarli, MD (University of California, Davis Sacramento); Smita Kittur, MD (Neurological Care of CNY); Michael Borrie, MB ChB (Parkwood Hospital); T Y Lee, PhD (Parkwood Hospital); Dr Rob Bartha, PhD (Parkwood Hospital); Sterling Johnson, PhD (University of Wisconsin); Sanjay Asthana, MD (University of Wisconsin); Cynthia M. Carlsson, MD (University of Wisconsin); Steven G. Potkin, MD (University of California, Irvine BIC); Adrian Preda, MD (University of California, Irvine BIC); Dana Nguyen, PhD (University of California, Irvine BIC); Pierre Tariot, MD (Banner Alzheimer’s Institute); Adam Fleisher, MD (Banner Alzheimer’s Institute); Stephanie Reeder, BA (Banner Alzheimer’s Institute); Vernice Bates, MD (Dent Neurologic Institute); Horacio Capote, MD (Dent Neurologic Institute); Michelle Rainka, PharmD, CCRP (Dent Neurologic Institute); Douglas W. Scharre, MD (Ohio State University); Maria Kataki, MD, PhD (Ohio State University); Anahita Adeli, MD (Ohio State University); Earl A. Zimmerman, MD (Albany Medical College); Dzintra Celmins, MD (Albany Medical College); Alice D. Brown, FNP (Albany Medical College); Godfrey D. Pearlson, MD (Hartford Hosp, Olin Neuropsychiatry Research Center); Karen Blank, MD (Hartford Hosp, Olin Neuropsychiatry Research Center); Karen Anderson, RN (Hartford Hosp, Olin Neuropsychiatry Research Center); Robert B. Santulli, MD (Dartmouth Hitchcock Medical Center); Tamar J. Kitzmiller (Dartmouth Hitchcock Medical Center); Eben S. Schwartz, PhD (Dartmouth Hitchcock Medical Center); Kaycee M. Sink, MD, MAS (Wake Forest University Health Sciences); Jeff D. Williamson, MD, MHS (Wake Forest University Health Sciences); Pradeep Garg, PhD (Wake Forest University Health Sciences); Franklin Watkins, MD (Wake Forest University Health Sciences); Brian R. Ott, MD (Rhode Island Hospital); Henry Querfurth, MD (Rhode Island Hospital); Geoffrey Tremont, PhD (Rhode Island Groups Acknowledgements Journal Format 3 Hospital); Stephen Salloway, MD, MS (Butler Hospital); Paul Malloy, PhD (Butler Hospital); Stephen Correia, PhD (Butler Hospital); Howard J. Rosen, MD (UC San Francisco); Bruce L. Miller, MD (UC San Francisco); Jacobo Mintzer, MD, MBA (Medical University South Carolina); Kenneth Spicer, MD, PhD (Medical University South Carolina); David Bachman, MD (Medical University South Carolina); Elizabether Finger, MD (St. Joseph’s Health Care); Stephen Pasternak, MD (St. Joseph’s Health Care); Irina Rachinsky, MD (St. Joseph’s Health Care); John Rogers, MD (St. Joseph’s Health Care); Andrew Kertesz, MD (St. Joseph’s Health Care); Dick Drost, MD (St. Joseph’s Health Care); Nunzio Pomara, MD (Nathan Kline Institute); Raymundo Hernando, MD (Nathan Kline Institute); Antero Sarrael, MD (Nathan Kline Institute); Susan K. Schultz, MD (University of Iowa College of Medicine, Iowa City); Laura L. Boles Ponto, PhD (University of Iowa College of Medicine, Iowa City); Hyungsub Shim, MD (University of Iowa College of Medicine, Iowa City); Karen Elizabeth Smith, RN (University of Iowa College of Medicine, Iowa City); Norman Relkin, MD, PhD (Cornell University); Gloria Chaing, MD (Cornell University); Lisa Raudin, PhD (Cornell University); Amanda Smith, MD (University of South Floriday: USF Health Byrd Alzheimer’s Institute); Kristin Fargher, MD (University of South Floriday: USF Health Byrd Alzheimer’s Institute); Balebail Ashok Raj, MD (University of South Floriday: USF Health Byrd Alzheimer’s Institute)

## REFERENCES

[1] Tagariello P, Girardi P, Amore M. Depression and apathy in dementia: same syndrome or different constructs? A critical review. Arch Gerontol Geriatr. 2009;49:246–9.

[2] Leung DKY, Chan WC, Spector A, Wong GHY. Prevalence of depression, anxiety, and apathy symptoms across dementia stages: A systematic review and meta-analysis. International journal of geriatric psychiatry. 2021;36:1330–44.

[3] Lanctot KL, Amatniek J, Ancoli-Israel S, Arnold SE, Ballard C, Cohen-Mansfield J, et al. Neuropsychiatric signs and symptoms of Alzheimer’s disease: New treatment paradigms. Alzheimers Dement (N Y). 2017;3:440–9.

[4] van Reekum R, Stuss DT, Ostrander LJTJon, neurosciences c. Apathy: why care? 2005;17:7–19.

[5] Marin Rsjpa. Differential diagnosis of apathy and related disorders of diminished motivation. 1997;27:30–3.

[6] Marin RSJTJon, neurosciences c. Apathy: a neuropsychiatric syndrome. 1991.

[7] Starkstein SE, Petracca G, Chemerinski E, Kremer J. Syndromic validity of apathy in Alzheimer’s disease. Am J Psychiatry. 2001;158:872–7.

[8] Rajkumar AP, Ballard C, Fossey J, Corbett A, Woods B, Orrell M, et al. Apathy and Its Response to Antipsychotic Review and Nonpharmacological Interventions in People With Dementia Living in Nursing Homes: WHELD, a Factorial Cluster Randomized Controlled Trial. J Am Med Dir Assoc. 2016;17:741–7.

[9] Ishii S, Weintraub N, Mervis JR. Apathy: a common psychiatric syndrome in the elderly. J Am Med Dir Assoc. 2009;10:381–93.

[10] Lanctot KL, Aguera-Ortiz L, Brodaty H, Francis PT, Geda YE, Ismail Z, et al. Apathy associated with neurocognitive disorders: Recent progress and future directions. Alzheimers Dement. 2017;13:84–100.

[11] Benoit M, Berrut G, Doussaint J, Bakchine S, Bonin-Guillaume S, Fremont P, et al. Apathy and depression in mild Alzheimer’s disease: a cross-sectional study using diagnostic criteria. J Alzheimers Dis. 2012;31:325–34.

[12] Selbaek G, Engedal K, Bergh S. The prevalence and course of neuropsychiatric symptoms in nursing home patients with dementia: a systematic review. J Am Med Dir Assoc. 2013;14:161–9.

[13] Miller DS, Robert P, Ereshefsky L, Adler L, Bateman D, Cummings J, et al. Diagnostic criteria for apathy in neurocognitive disorders. Alzheimer’s & Dementia. 2021;17:1892–904.

[14] Cummings J. The Neuropsychiatric Inventory: Development and Applications. Journal of Geriatric Psychiatry and Neurology. 2020;33:73–84.

[15] Cummings JL. The Neuropsychiatric Inventory: assessing psychopathology in dementia patients. Neurology. 1997;48:S10–6.

[16] Wood S, Cummings JL, Hsu M-A, Barclay T, Wheatley MV, Yarema KT, et al. The Use of the Neuropsychiatric Inventory in Nursing Home Residents: Characterization and Measurement. The American Journal of Geriatric Psychiatry. 2000;8:75–83.

[17] Morris JC, McKeel Jr DW, Fulling K, Torack RM, Berg L. Validation of clinical diagnostic criteria for Alzheimer’s disease. Annals of Neurology. 1988;24:17–22.

[18] Lanza ST, Dziak JJ, Huang L, Wagner AT, Collins LM. LCA Stata Plugin Users’ Guide Version 1.2.1. University Park: The Methodology Center, Penn State. 2018.

[19] Huang L, Dziak JJ, Wagner AT, Lanza ST. LCA Bootstrap Stata function users’ guide (Version 1.0). University park: the methodology center, Penn State. 2016.

[20] Nylund KL, Asparouhov T, Muthén BO. Deciding on the Number of Classes in Latent Class Analysis and Growth Mixture Modeling: A Monte Carlo Simulation Study. Structural Equation Modeling: A Multidisciplinary Journal. 2007;14:535–69.

[21] Huang L, Dziak JJ, Bray BC, Wagner AT. LCA_Distal_BCH Stata function Users’ Guide (Version 1.1). 2017.

[22] Bolck A, Croon M, Hagenaars J. Estimating latent structure models with categorical variables: One-step versus three-step estimators. Political analysis. 2004;12:3–27.

[23] Vermunt JK. Latent class modeling with covariates: Two improved three-step approaches. Political analysis. 2010;18:450–69.

[24] Clarke DE, van Reekum R, Simard M, Streiner DL, Conn D, Cohen T, et al. Apathy in Dementia: Clinical and Sociodemographic Correlates. The Journal of neuropsychiatry and clinical neurosciences. 2008;20:337–47.

[25] Fischer CE, Ismail Z, Youakim JM, Creese B, Kumar S, Nuñez N, et al. Revisiting Criteria for Psychosis in Alzheimer’s Disease and Related Dementias: Toward Better Phenotypic Classification and Biomarker Research. Journal of Alzheimer’s Disease. 2020;73:1143–56.

[26] Cummings J, Pinto LC, Cruz M, Fischer CE, Gerritsen DL, Grossberg GT, et al. Criteria for Psychosis in Major and Mild Neurocognitive Disorders: International Psychogeriatric Association (IPA) Consensus Clinical and Research Definition. The American journal of geriatric psychiatry : official journal of the American Association for Geriatric Psychiatry. 2020;28:1256–69.

[27] Sweet RA, Bennett DA, Graff-Radford NR, Mayeux R. Assessment and familial aggregation of psychosis in Alzheimer’s disease from the National Institute on Aging Late Onset Alzheimer’s Disease Family Study. Brain. 2010;133:1155–62.

[28] Lyketsos CG, Sheppard JM, Steinberg M, Tschanz JA, Norton MC, Steffens DC, et al. Neuropsychiatric disturbance in Alzheimer’s disease clusters into three groups: the Cache County study. International journal of geriatric psychiatry. 2001;16:1043–53.

[29] Wilkosz PA, Miyahara S, Lopez OL, Dekosky ST, Sweet RA. Prediction of psychosis onset in Alzheimer disease: The role of cognitive impairment, depressive symptoms, and further evidence for psychosis subtypes. The American journal of geriatric psychiatry : official journal of the American Association for Geriatric Psychiatry. 2006;14:352–60.

[30] Ismail Z, Creese B, Aarsland D, Kales HC, Lyketsos CG, Sweet RA, et al. Psychosis in Alzheimer disease — mechanisms, genetics and therapeutic opportunities. Nature Reviews Neurology. 2022;18:131–44.

[31] DeMichele-Sweet MAA, Klei L, Creese B, Harwood JC, Weamer EA, McClain L, et al. Genome-wide association identifies the first risk loci for psychosis in Alzheimer disease. Molecular Psychiatry. 2021;26:5797–811.

[32] Stringaris A, Zavos H, Leibenluft E, Maughan B, Eley TC. Adolescent irritability: phenotypic associations and genetic links with depressed mood. The American journal of psychiatry. 2012;169:47–54.

[33] Agüera-Ortiz L, Babulal GM, Bruneau M-A, Creese B, D’Antonio F, Fischer CE, et al. Psychosis as a Treatment Target in Dementia: A Roadmap for Designing Interventions. Journal of Alzheimer’s Disease. 2022;Preprint:1–26.

[34] Anatchkova M, Brooks A, Swett L, Hartry A, Duffy RA, Baker RA, et al. Agitation in patients with dementia: a systematic review of epidemiology and association with severity and course. International Psychogeriatrics. 2019;31:1305–18.

[35] de Medeiros K, Robert P, Gauthier S, Stella F, Politis A, Leoutsakos J, et al. The Neuropsychiatric Inventory-Clinician rating scale (NPI-C): reliability and validity of a revised assessment of neuropsychiatric symptoms in dementia. International Psychogeriatrics. 2010;22:984–94.

